# Multi-omics Mendelian randomisation using expression, splicing and protein quantitative trait loci: identification of novel drug targets for gliomagenesis

**DOI:** 10.1101/2022.05.11.22274959

**Authors:** Zak A Thornton, Lily J Andrews, Huiling Zhao, Jie Zheng, Lavinia Paternoster, Jamie W Robinson, Kathreena M Kurian

## Abstract

**Background:** Genetic variants associated with molecular traits that are also associated with liability to glioma can provide causal evidence for the prioritisation of these traits as candidate drug targets.

**Methods:** We performed two-sample Mendelian randomisation and genetic colocalisation of a large panel of molecular traits on glioma. Molecular data were taken from studies of expression quantitative trait loci (QTL) [11,985 genes]; splicing QTL [13,285 genes] and protein QTL [7,376 proteins] derived from 15 brain tissues. Glioma summary-level data was extracted from a genome-wide association meta-analysis of 12,496 cases and 18,190 controls.

**Results:** Our MR analysis showed evidence for a causal effect of 85 molecular traits on glioma – 37 were robust according to colocalisation and Steiger filtering. We found causal evidence for 10 genes previously associated with glioma risk. We identified one novel genetic locus with strong causal evidence in the gene expression analysis: *HBEGF* (5q31.3) in all glioma [OR 1.36 (95%CI 1.19 to 1.55); P = 4.41 x 10^-6^]. We also identified three novel genetic loci with strong causal evidence in the splicing variation analysis: *CEP192* (18p11.21) in glioblastoma [OR 4.40 (95%CI 2.28 to 8.48); P = 9.78 x 10^-4^]; *FAIM* (3q22.3) in all glioma [OR 2.72 to 3.43; P = 1.03 x 10^-5^ to 1.09 x 10^-5^] and *SLC8A1* (2p22.1) in all glioma [OR 0.37 (95%CI 0.24 to 0.56; P = 5.72 x 10^-6^].

**Conclusions:** We provide robust causal evidence for genes previously associated with glioma risk in genome-wide association studies, as well as four novel genes.

## Introduction

Glioma is the most common (∼80%) primary malignant brain and central nervous system (CNS) tumour group, with age-standardised incidence rates ranging from 4.67 to 5.73 per 100,000. ^1,2^ Malignant brain tumours are responsible for the greatest years of potential life lost of all cancers (∼20 years), with a comparatively shorter 5-year relative survival rate (34.9%) compared to other malignancies. ^3,4^

In the 2016 World Health Organisation (WHO) guidelines for diagnosis of brain tumours, histological markers formed a sufficient basis to classify CNS tumours; however, this was expanded in the latest 2021 edition, which suggests molecular profiling as a means of stratifying tumour types. ^5,6^ The reason for this change is to emphasise the shift in how glioma, and brain tumours in general, are now classified in the clinic. One key example is the use of *MGMT* methylation as a tool to stratify patients in clinical trials. ^7,8^ However, the molecular landscape of glioma remains complex and heterogeneous, highlighting an area of unmet research need to improve diagnosis, patient stratification and, hopefully, treatment.

One approach to elucidating novel molecular markers comes from the field of genetic epidemiology in the way of genome-wide association studies (GWAS) and transcriptome-wide association studies (TWAS), two study designs which leverage large-scale human genotyping of genetic variation to determine which genes are associated with a phenotype of interest. To date, previous studies have identified 27 genetic loci associated with glioma risk, implicating a total of approximately 50 genes; however, further analysis is required to determine both which genes these genetic variants map to, and whether such variation is causally implicated in gliomagenesis, or simply an observational association. ^9–14^

Furthermore, genotyping of large cohorts has been performed to link disease-associated genetic variants to gene regulatory mechanisms to improve understanding of disease aetiology. ^15^ Such genetic variants associated with these molecular traits are known as quantitative trait loci (QTL), and examples of these include measuring relative gene expression levels (eQTLs), splicing variation (sQTLs) and protein abundance (pQTLs). Integrating multi-dimensional omics data can further our understanding of the aetiology of complex traits by providing functional and mechanistic insight into these layered molecular relationships. ^16–18^

Mendelian randomisation (MR) is an established statistical method which utilises genetic predictors of an exposure to assess the causal relationship between this exposure and an outcome. ^19^ The main advantage MR has over traditional observational epidemiological methods is that MR can determine causality between an exposure and an outcome as it is less liable to common epidemiological biases, such as confounding and reverse causality. When combined with other sensitivity analyses such as genetic colocalisation, MR can provide strong evidence of an unbiased causal relationship between genes and traits of interest. ^20^

In this study, we leveraged multi-dimensional QTL data derived in bulk brain tissues in a combined MR-colocalisation framework, with the aim to identify causal evidence for aetiologically important genes for gliomagenesis.

## Methods

A diagram of the workflow described in this section can be found in Figure 1.

**Figure 1.**
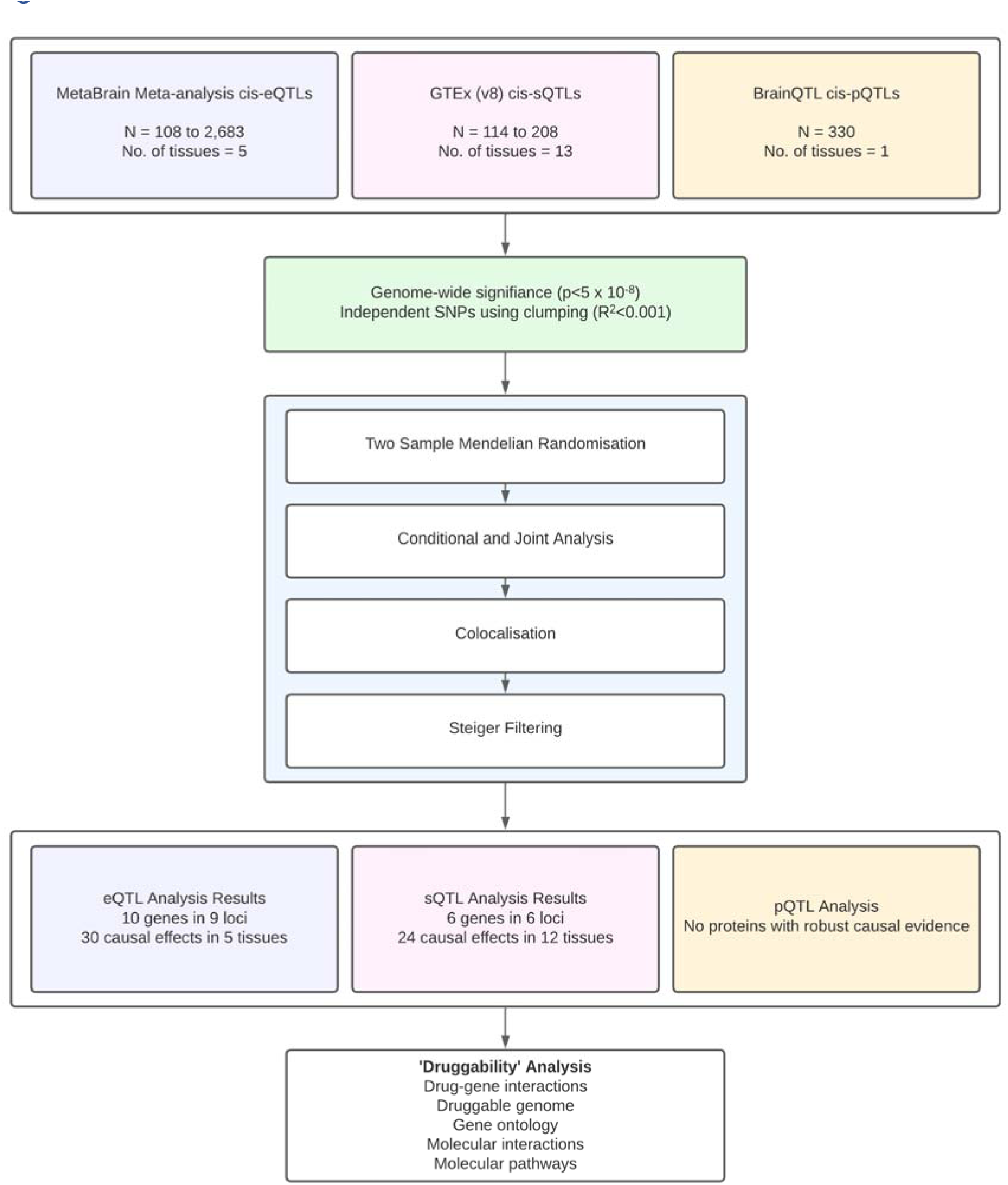
Flowchart showing the pipeline used in the multi-omic analysis.

### Study Datasets

We used QTL summary statistics from genome-wide association studies (GWAS) of gene expression, protein abundance and alternative splicing levels in various brain tissues described below. Overviews of these datasets are given in Supplementary Table S1.

Gene expression data were taken from MetaBrain, a meta-analysis of 14 eQTL datasets derived from 3,659 samples from 2,683 individuals of European ancestry in five CNS tissues: the basal ganglia (n=208), cerebellum (n=492), cortex (n=2,683), hippocampus (n=168), and spinal cord (n=108) (Supplementary Table S1). ^21^

The sQTL data were taken from GTEx Portal (v8 release). We used sGenes provided by GTEx, generated using their QC protocol, from a primarily European-American (85.3%) population. sQTL were extracted from 13 regions of the CNS: amygdala (n=129), anterior cingulate cortex (BA24) (n=147), caudate (n=194), cerebellar hemisphere (n=175), cerebellum (n=209), cortex (n=205), frontal cortex (BA9) (n=175), hippocampus (n=165), hypothalamus (n=170), nucleus accumbens (n=202), putamen (n=170), spinal cord (n=126), and substantia nigra (n=114) (Supplementary Table S1). ^22^

Finally, we included pQTL data retrieved from BrainQTL, derived from European ancestry individuals in the religious orders study and memory and ageing projects. ^23,24^ pQTL were extracted from the dorsolateral pre-frontal cortex of 330 individuals (Supplementary Table S1). ^25^

Glioma summary-level data were derived from a meta-analysis of eight constituent glioma GWAS consisted of 6,191 glioblastoma (GB) cases, 5,819 non-GB cases, 12,496 combined cases and 18,190 controls. ^9^ We therefore used three outcomes throughout our analyses: the GB-only case load, the non-GB-only case load, and the combined case load (defined as ‘all glioma’). Although glioma is highly heterogeneous, we did this to increase our statistical power by using larger sample sizes. The non-GB cases analysed in this meta-analysis were not explicitly classified but consists mainly of astrocytoma and oligodendroglioma diagnosed patients (Supplementary Table S1).

This study uses previously published summary-level data and thus contains no patient identifiable data. Ethical approval and informed consent from each participant were given and can be found where the dataset was initially described. All procedures performed in studies involving human participants were done in accordance with the ethical standards of the institutional or national research committee and with 1964 Helsinki declaration.

### Instrument Selection

We identified *cis-*acting (within 1Mb of the gene coding region) QTLs which met genome-wide significance (P < 5×10^-8^). *Trans-*acting QTLs were excluded from the analysis because of the increased likelihood of horizontal pleiotropy, due to their distant location from the gene whose variation they alter. Instruments were selected to be in linkage disequilibrium (R^2^ < 0.001) to ensure independence.

### Two Sample Mendelian Randomisation

We used a two-sample MR framework to estimate the causal effect of genetically proxied gene expression, protein abundance and alternative splicing on genetic liability to glioma risk. MR estimates were generated using the Wald ratio method for instruments consisting of single SNPs and inverse variance weighted (IVW) method for instruments comprising of multiple SNPs. ^26^ MR estimates were transformed and presented throughout as odds ratios (OR) and were scaled to reflect one standard deviation increase in the respective molecular trait. Following MR analysis, the results had to meet a Bonferroni-corrected P value threshold (0.05/number of tests performed) to adjust for multiple testing.

MR has three assumptions which must hold to produce an unbiased estimate. Firstly, the genetic instrument must associate with the exposure (‘relevance’). We tested this by generating the F-statistic for each instrument, where an F-statistic > 10 is evidence against weak instrument bias, and filtering out instruments which did not surpass this threshold. ^27,28^ The second assumption is that the genetic instrument does not share a common cause with the outcome (‘independence’). We tested this assumption partly by using genetic colocalisation, which determines whether the molecular trait and glioma share the same causal variant, a necessary (though not sufficient) condition for causality. Using colocalisation in this way has been posited to at least eliminate some unreliable associations when standard follow up sensitivity analyses to evaluate the presence of horizontal pleiotropy (such as MR-Egger) are unavailable. ^27,29^ The final assumption is that the genetic instrument affects the outcome only through the effect of the exposure of interest (‘exclusion restriction’). This assumption, also known as horizontal pleiotropy, is difficult to assess with single SNP instruments, as is common for ‘omic’ variables. This was tested by investigating whether there was causal evidence linking the molecular trait of interest with putative risk factors for glioma. The presence of an association between the molecular trait and glioma risk factors may then be evidence of potential horizontal pleiotropy (Figure 2). ^27^

**Figure 2.**
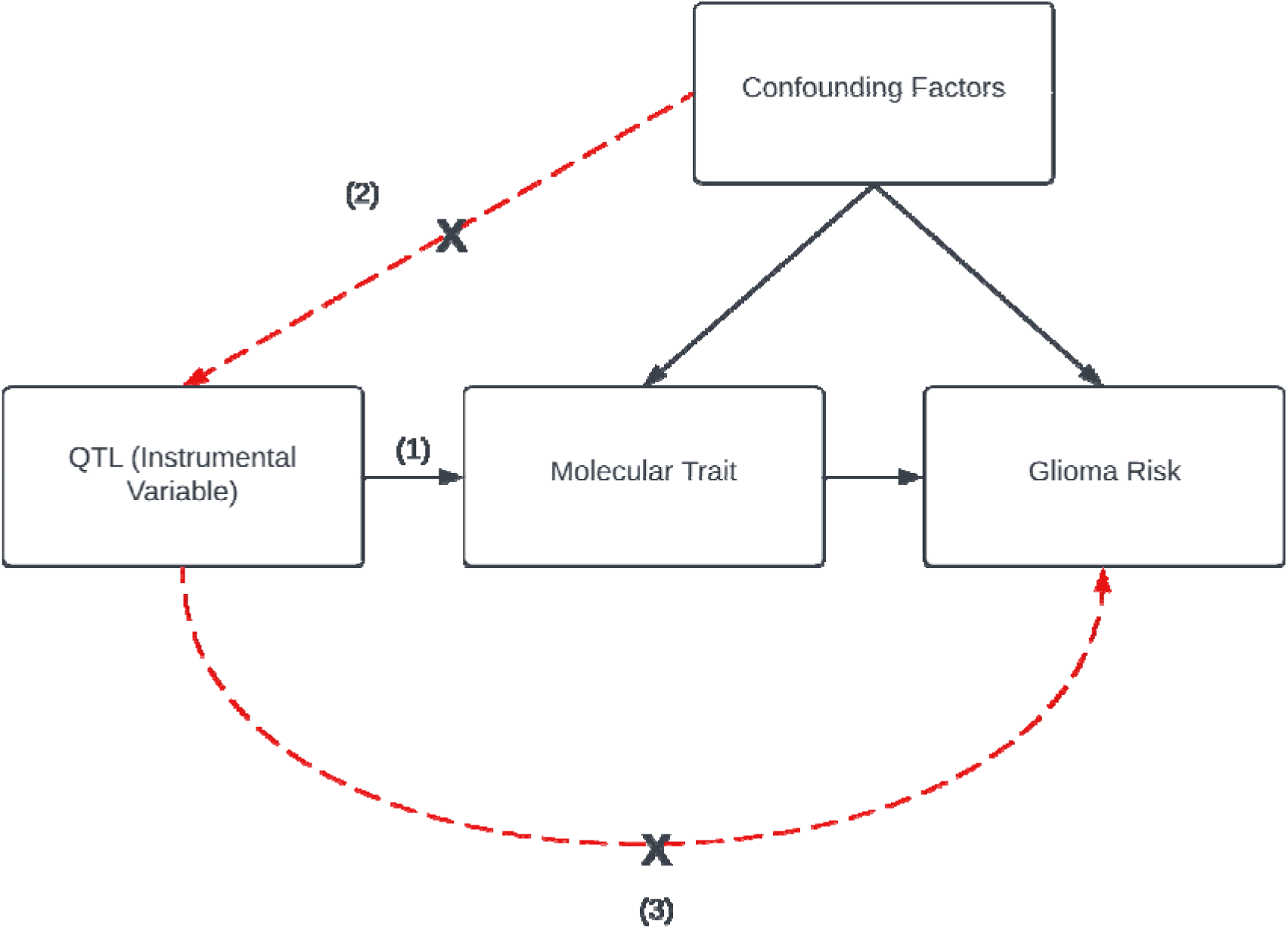
Directed Acyclic Graph (DAG) showing a visual representation of potential causal relationships. This DAG also shows the three assumptions of Mendelian randomisation. (1) The genetic variant(s) being used as an instrument is associated robustly with the exposure. (2) The instrument is independent of measured and unmeasured confounding factors of the association between exposure and outcome. (3) There must be no independent pathway between the instrument and outcome other than through the exposure.

For brevity, we refer to our MR results as causal genes (or causal proteins); however, we recognise that further studies and evidence will be required to validate our results.

### Colocalisation

We tested for colocalisation between the loci of each molecular trait and glioma subtype which had an MR result that passed the Bonferroni-corrected P value threshold. We extracted *cis-*regions consisting of all SNPs within a 1Mb window around the gene coding regions. ^20^ Colocalisation requires the *cis*-region of the molecular trait to be richly genotyped. If there were not enough SNPs to perform colocalisation, we used LDCheck to measure the pairwise LD between the QTL and *cis*-SNPs in the glioma data at a more lenient threshold (P < 5 x 10^-6^). If the SNPs were in high LD (R^2^ > 0.8), then we took this as proxy, albeit weak, evidence for colocalisation. This method has been described and used previously when full genome-wide statistics were not available. ^30^

### Steiger Filtering

For causal effects which had strong evidence of colocalisation, we applied Steiger filtering to test for the presence of reverse causation, where the instrument explains more of the variance in the outcome than the exposure.^29^ Results of Steiger filtering are presented as a one of three categorical variables; “pass” if Steiger P < 0.05 and the instrument explains more of the variance in the outcome than the exposure, “fail” if Steiger P < 0.05 and the instrument explains more of the variance in the exposure than the outcome, and “uncertain” if P ≥ 0.05.

### Evidence thresholds for multi-omic analyses

Our main results comprised of those which passed the Bonferroni correction P value, had strong colocalisation evidence (H_4_ ≥ 80%) and passed Steiger filtering sensitivity analysis, as they have the most robust evidence. Since Steiger filtering is sensitive to sample sizes, the analysis may be liable to false negatives if one dataset is better powered than the other; therefore, given the relatively small samples in the QTL datasets, we included results which had “uncertain” evidence but with the caveat that these results may be less robust.

### Overlap in signals between molecular traits

To determine whether there was any potential overlap in signals between molecular traits, we looked at all MR results for our molecular traits with robust evidence in all tissues, regardless of P value. We then filtered these results using a more lenient Bonferroni correction P value (similarly, by using 0.05/number of tests performed) to determine whether any were potentially overlapping signals which failed to pass due to insufficient statistical power.

### Specificity Analysis

To annotate the potential causal pathway between the molecular trait and glioma, we used two different approaches: exploring PheWAS data, and risk factors with pre-existing evidence in the literature. To perform a broad search for potential causal pathways, we used the IEU OpenGWAS project PheWAS tool and PhenoScanner V2 to search for phenotypes which were strongly associated (P < 5 x 10^-8^) with lead SNPs used to instrument the robust causal effects. ^31–33^ We then performed a specificity analysis to determine whether the robust causal effects were being caused by non-specific effects or driven through a putative risk factor for glioma. We investigated risk factors with genetic liability to allergies, asthma, altered low-density lipoprotein cholesterol (LDLc) levels and alcohol consumption. These risk factors were selected based on potential evidence linking that risk factor to glioma in the literature, if there was data available for that risk factor in the IEU OpenGWAS project database, and whether those data were available for use in an MR analysis. ^34^

### Differential expression in molecular traits

We explored whether there was differential expression in the genes for which there was robust evidence that expression had a causal effect on at least one glioma subtype, between subtypes of glioma and control patients using the GlioVis resource. ^35^

We investigated the differences in the expression of splicing variants in tissue types in the genes for which there was evidence that gene-splicing had a causal effect on at least one glioma subtype, using the GTEx portal. ^22^

### Annotation of ‘druggability’

We annotated robust results which passed sensitivity analyses with evidence of ‘druggability’, i.e. how likely a gene and its gene product are to be a valid drug target, to build evidence for the prioritisation of genes and proteins as candidate targets. We used the Open Targets platform, a large-scale database that uses genetic and genomic data for systematic drug target identification and prioritisation, to search for gene ontology terms, pathways, and gene interactions. ^36,37^ We also used Drug Gene Interaction Database (DGIdb) to search for drug-gene interactions and genome ‘druggability’. ^38^

### Software

We used the TwoSampleMR R package to implement the MR analyses and Steiger filtering. ^39–41^ We performed genetic colocalisation using Pair Wise Conditional Analysis and Colocalisation (PWCoCo), a method which combines Conditional and Joint Analysis (GCTA-COJO) and colocalisation (Coloc R package) to ensure the single causal variant assumption of colocalisation holds. ^20,42,43^

## Results

### Estimating the causal effects of gene expression on glioma risk

Of the 10,488 genes for which expression could be instrumented, 10,006 (95.4%) had the relevant SNPs available in the glioma GWAS and therefore MR could be performed. We found 29 genes with evidence for a causal effect (P < 5.00 x 10^-6^) on all glioma, GB, or non-GB in at least one of the tissues tested, resulting in 80 causal effects in total; 40 causal effects in cortex, 30 in cerebellum, 6 in basal ganglia, 3 in hippocampus and 1 in spinal cord (Supplementary Figure S1). In the follow-up sensitivity analyses of the unique 80 expression causal effects, we found that 30 (38%) had strong evidence of colocalisation (H_4_ > 80%), 11 (14%) had moderate evidence of colocalisation (80% > H_4_ > 50%) and 39 (49%) had weak evidence of colocalisation (H_4_ < 50%). The 30 causal effects (in 10 genes) with strong evidence for colocalisation were then tested for the potential of reverse causality using Steiger filtering; 29 causal effects passed this test, whilst one result was uncertain according to Steiger P-value (P > 0.05, Table 1).

**Table 1.** Results of the MR analysis investigating the effects of genetically proxied gene expression levels on genetic liability to glioma subtype.

| Tissue | Gene | Subtype | Method | No. of SNPs | P Value | Odds Ratio (95% CI) | H4 (%) | Steiger Direction | Steiger P Value | Steiger Flag |
| --- | --- | --- | --- | --- | --- | --- | --- | --- | --- | --- |
| Cortex | *CDKN2B* | All | Wald ratio | 1 | 7.17E-43 | 1.36 (1.19 - 1.55) | 93 | TRUE | 2.32E-19 | TRUE |
| Cortex | *CDKN2B* | GB | Wald ratio | 1 | 6.22E-44 | 1.14 (1.08 - 1.21) | 97 | TRUE | 3.19E-15 | TRUE |
| Cortex | *CDKN2B* | Non-GB | Wald ratio | 1 | 1.59E-14 | 1.22 (1.15 - 1.29) | 92 | TRUE | 2.91E-21 | TRUE |
| Cerebellum | *EGFR* | All | Wald ratio | 1 | 1.07E-25 | 1.22 (1.14 - 1.32) | 96 | TRUE | 2.16E-10 | TRUE |
| Cerebellum | *EGFR* | GB | Wald ratio | 1 | 2.89E-31 | 0.67 (0.63 - 0.73) | 97 | TRUE | 4.29E-09 | TRUE |
| Cerebellum | *EGFR* | Non-GB | Wald ratio | 1 | 1.60E-07 | 0.58 (0.53 - 0.64) | 98 | TRUE | 2.65E-11 | TRUE |
| Basal Ganglia | *GALNT6* | All | Wald ratio | 1 | 3.14E-06 | 0.5 (0.46 - 0.55) | 100 | TRUE | 4.78E-09 | TRUE |
| Basal Ganglia | *GALNT6* | GB | Wald ratio | 1 | 9.08E-08 | 0.43 (0.38 - 0.48) | 100 | TRUE | 1.23E-08 | TRUE |
| Cerebellum | *GALNT6* | All | Wald ratio | 1 | 3.14E-06 | 1.2 (1.12 - 1.28) | 100 | TRUE | 7.94E-25 | TRUE |
| Cerebellum | *GALNT6* | GB | Wald ratio | 1 | 9.08E-08 | 1.2 (1.12 - 1.29) | 100 | TRUE | 1.76E-23 | TRUE |
| Cortex | *GALNT6* | GB | Inverse variance weighted | 2 | 1.53E-07 | 1.19 (1.13 - 1.25) | 100 | TRUE | 6.87E-130 | TRUE |
| Hippocampus | *GALNT6* | All | Wald ratio | 1 | 4.17E-06 | 6.89 (5.60 - 8.47) | 99 | TRUE | 5.29E-08 | TRUE |
| Hippocampus | *GALNT6* | GB | Wald ratio | 1 | 2.15E-07 | 1.87 (1.50 - 2.34) | 100 | TRUE | 1.09E-07 | TRUE |
| Spinal Cord | *GALNT6* | GB | Wald ratio | 1 | 8.40E-07 | 2.71 (2.16 - 3.40) | 99 | TRUE | 6.88E-07 | TRUE |
| Cortex | *HBEGF* | All | Wald ratio | 1 | 4.41E-06 | 1.17 (1.12 - 1.23) | 99 | TRUE | 8.18E-17 | TRUE |
| Basal Ganglia | *HEATR3* | All | Wald ratio | 1 | 1.50E-10 | 0.77 (0.69 - 0.85) | 93 | TRUE | 7.61E-11 | TRUE |
| Basal Ganglia | *HEATR3* | GB | Wald ratio | 1 | 1.55E-11 | 0.60 (0.53 - 0.68) | 95 | TRUE | 2.58E-11 | TRUE |
| Cerebellum | *HEATR3* | All | Wald ratio | 1 | 3.93E-11 | 1.15 (1.09 - 1.23) | 96 | TRUE | 6.45E-54 | TRUE |
| Cerebellum | *HEATR3* | GB | Wald ratio | 1 | 6.74E-12 | 1.17 (1.11 - 1.24) | 97 | TRUE | 1.25E-51 | TRUE |
| Cortex | *HEATR3* | All | Wald ratio | 1 | 1.95E-10 | 1.26 (1.18 - 1.34) | 97 | TRUE | 1.15E-139 | TRUE |
| Cortex | *HEATR3* | GB | Wald ratio | 1 | 8.20E-11 | 1.14 (1.1 - 1.19) | 97 | TRUE | 3.76E-125 | TRUE |
| Cerebellum | *JAK1* | GB | Wald ratio | 1 | 9.28E-09 | 1.14 (1.08 - 1.20) | 89 | TRUE | 2.68E-38 | TRUE |
| Cerebellum | *MDM4* | All | Wald ratio | 1 | 9.54E-07 | 1.18 (1.11 - 1.26) | 91 | TRUE | 2.10E-08 | TRUE |
| Cortex | *PICK1* | GB | Wald ratio | 1 | 1.92E-09 | 1.19 (1.13 - 1.26) | 91 | TRUE | 2.14E-31 | TRUE |
| Hippocampus | *PICK1* | GB | Wald ratio | 1 | 3.95E-09 | 4.43 (3.74 - 5.25) | 81 | TRUE | 1.65E-06 | TRUE |
| Cortex | *RAVER2* | All | Wald ratio | 1 | 3.19E-08 | 0.80 (0.73 - 0.87) | 94 | TRUE | 8.11E-06 | TRUE |
| Cortex | *RAVER2* | GB | Wald ratio | 1 | 7.92E-10 | 0.86 (0.81 - 0.90) | 98 | TRUE | 1.42E-04 | TRUE |
| Cortex | *TERT* | All | Wald ratio | 1 | 2.32E-66 | 1.27 (1.17 - 1.37) | 100 | TRUE | 3.55E-03 | TRUE |
| Cortex | *TERT* | GB | Wald ratio | 1 | 6.92E-75 | 1.39 (1.25 - 1.55) | 100 | TRUE | 1.18E-01 | UNCERTAIN |
| Cortex | *TERT* | Non-GB | Wald ratio | 1 | 6.69E-18 | 2.35 (1.79 - 3.08) | 100 | TRUE | 3.26E-05 | TRUE |

*Results presented are those which were deemed the most robust, i.e., met the Bonferroni-correct P value threshold ( $P = 5.00 \times 10^{-6}$ ) for the MR analysis, had* *strong evidence of colocalisation ( $H4 > 80\%$ ) and correctly orientated Steiger filtering direction.*

Three genes (*GALNT6, HEATR3, PICK1*) with robust evidence of a causal effect were identified in multiple tissues, with different lead SNPs, but all SNPs were in high LD (R^2^ > 0.7). ^30^ Two genes (*JAK1* and *PICK1*) with robust evidence had a causal effect in only one subtype (non-GB and GB, respectively) whereas all other genes (CDKN2B, *EGFR, GALNT6, HBEGF, HEATR3, MDM4, RAVER2* and *TERT*) had a causal effect on all glioma, or more than one outcome tested (Table 1).

In summary, we found 10 genes for which there was robust evidence that expression had a causal effect on at least one glioma subtype in at least one brain region (30 causal effects in total, Figure 3). Three genes (*CDKN2B, RAVER2, TERT*) had strong causal effects (OR <0.5 or >2). For six genes, increased expression was causally related to increased odds of glioma. All results for all eQTL sensitivity analyses are described in Supplementary Table S1.

**Figure 3.**
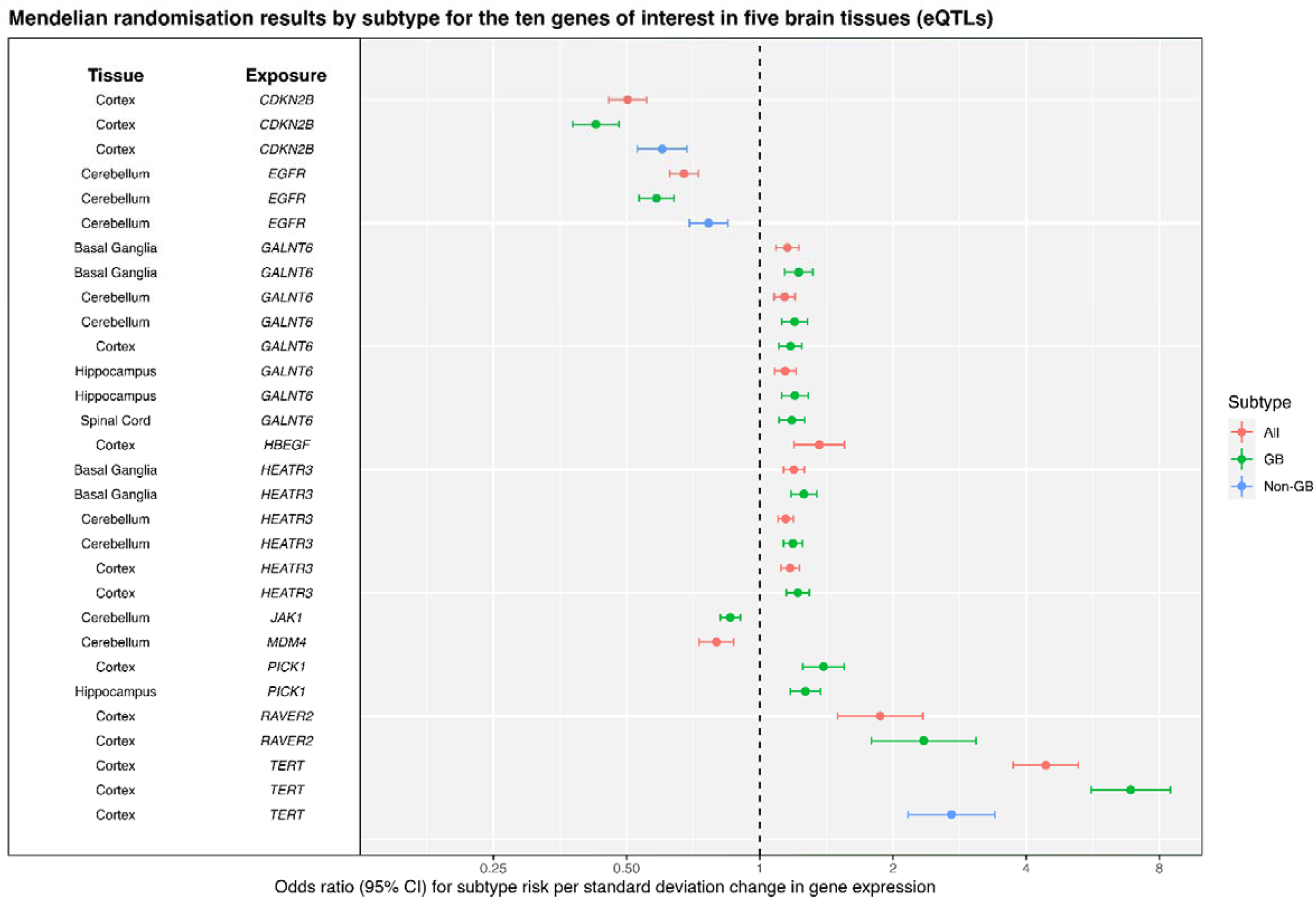
Forest plot showing Mendelian randomisation results by subtype (all glioma, GB and non-GB) for the ten genes of interest in five brain tissues in the gene expression analysis. Results have strong evidence of colocalization (H_4_ >= 80%) and correct Steiger directionality.

### Estimating the causal effects of alternative splicing on glioma risk

Of the 6,200 genes for which alternative splicing could be instrumented, 4,496 (72.5%) had the relevant SNPs available in the glioma GWAS are therefore MR could be performed. We found 10 genes with evidence for a causal effect (P < 1.11 x 10^-5^) on glioma, GB or non-GB in at least one of the thirteen tissues tested, resulting in 71 causal effects in total; five in amygdala, four in anterior cingulate cortex, seven in caudate, six in cerebellar hemisphere, nine in cerebellum, six in cortex, four in frontal cortex, seven in hippocampus, seven in hypothalamus, nine in nucleus accumbens, six in putamen and one in substantia nigra (Supplementary Figure S2). We applied the same sensitivity analysis pipeline. We found that 47 (66%) had strong evidence of colocalisation (H_4_ > 80%), five (7%) had moderate evidence of colocalisation (80% > H_4_ > 50%) and 19 (27%) had weak evidence of colocalisation (H_4_ < 50%). The 47 causal effects with strong evidence for colocalisation were then tested for the potential of reverse causality using Steiger filtering; 24 (51%) passed this test, however all of these results were uncertain according to Steiger P-value (P > 0.05. Table 2).

**Table 2.** Results of the MR analysis investigating the effects of genetically proxied gene splicing levels on genetic liability to glioma subtype.

| Tissue | Gene (Splice Junction) | Subtype | Method | No. of SNPs | P Value | Odds Ratio (95% CI) | H4 (%) | Steiger Direction | Steiger P Value | Steiger Flag |
| --- | --- | --- | --- | --- | --- | --- | --- | --- | --- | --- |
| Amygdala | *FAIM Splice Junction 5* | All | Wald ratio | 1 | 1.03E-05 | 2.72 (1.74 - 4.25) | 97 | TRUE | 6.91E-01 | UNCERTAIN |
| Amygdala | *RTEL1 Splice Junction 32* | All | Wald ratio | 1 | 4.38E-40 | 12.83 (8.80 - 18.72) | 100 | TRUE | 9.68E-01 | UNCERTAIN |
| Amygdala | *RTEL1 Splice Junction 32* | Non-GB | Wald ratio | 1 | 8.36E-10 | 4.61 (2.83 - 7.51) | 98 | TRUE | 7.34E-01 | UNCERTAIN |
| Anterior Cingulate Cortex | *RTEL1 Splice Junction 32* | All | Wald ratio | 1 | 3.89E-33 | 0.09 (0.06 - 0.13) | 94 | TRUE | 9.46E-01 | UNCERTAIN |
| Anterior Cingulate Cortex | *RTEL1 Splice Junction 32* | Non-GB | Wald ratio | 1 | 2.39E-08 | 0.23 (0.14 - 0.39) | 92 | TRUE | 7.24E-01 | UNCERTAIN |
| Caudate | *FAIM Splice Junction 5* | All | Wald ratio | 1 | 1.09E-05 | 3.01 (1.84 - 4.92) | 97 | TRUE | 6.83E-01 | UNCERTAIN |
| Caudate | *RTEL1 Splice Junction 32* | Non-GB | Wald ratio | 1 | 1.56E-10 | 7.13 (3.91 - 13.02) | 100 | TRUE | 8.68E-01 | UNCERTAIN |
| Cerebellar Hemisphere | *CEP192 Splice Junction 60* | GB | Wald ratio | 1 | 9.78E-06 | 4.40 (2.28 - 8.48) | 97 | TRUE | 7.79E-01 | UNCERTAIN |
| Cerebellar Hemisphere | *RTEL1 Splice Junction 32* | GB | Wald ratio | 1 | 1.58E-45 | 19.1 (12.7 - 28.72) | 100 | TRUE | 7.79E-01 | UNCERTAIN |
| Cerebellar Hemisphere | *RTEL1 Splice Junction 32* | All | Wald ratio | 1 | 4.38E-40 | 9.52 (6.82 - 13.28) | 100 | TRUE | 8.87E-01 | UNCERTAIN |
| Cerebellar Hemisphere | *RTEL1 Splice Junction 32* | Non-GB | Wald ratio | 1 | 8.36E-10 | 3.85 (2.51 - 5.93) | 95 | TRUE | 6.23E-01 | UNCERTAIN |
| Cerebellum | *HEATR3 Splice Junction 1* | All | Wald ratio | 1 | 1.29E-10 | 9.64 (4.83 - 19.24) | 91 | TRUE | 9.38E-01 | UNCERTAIN |
| Cerebellum | *RTEL1 Splice Junction 32* | Non-GB | Wald ratio | 1 | 2.32E-10 | 5.21 (3.13 - 8.68) | 99 | TRUE | 7.43E-01 | UNCERTAIN |
| Cortex | *RTEL1 Splice Junction 32* | GB | Wald ratio | 1 | 4.42E-47 | 52.42 (30.59 - 89.81) | 100 | TRUE | 6.97E-01 | UNCERTAIN |
| Frontal Cortex | *RTEL1 Splice Junction 32* | Non-GB | Wald ratio | 1 | 2.00E-10 | 5.28 (3.16 - 8.82) | 99 | TRUE | 7.94E-01 | UNCERTAIN |
| Hippocampus | *FAIM Splice Junction 5* | All | Wald ratio | 1 | 1.09E-05 | 2.76 (1.76 - 4.34) | 97 | TRUE | 6.64E-01 | UNCERTAIN |
| Hippocampus | *PHLDB1 Splice Junction 5* | All | Wald ratio | 1 | 7.83E-09 | 0.11 (0.05 - 0.24) | 87 | TRUE | 9.29E-01 | UNCERTAIN |
| Hippocampus | *RTEL1 Splice Junction 32* | Non-GB | Wald ratio | 1 | 8.36E-10 | 5.05 (3.01 - 8.46) | 96 | TRUE | 8.08E-01 | UNCERTAIN |
| Hypothalamus | *PHLDB1 Splice Junction 3* | All | Wald ratio | 1 | 8.40E-15 | 13.29 (6.92 - 25.55) | 99 | TRUE | 9.94E-01 | UNCERTAIN |
| Hypothalamus | *RTEL1 Splice Junction 32* | Non-GB | Wald ratio | 1 | 2.00E-10 | 6.02 (3.46 - 10.47) | 99 | TRUE | 8.12E-01 | UNCERTAIN |
| Nucleus Accumbens | *FAIM Splice Junction 5* | All | Wald ratio | 1 | 1.03E-05 | 3.43 (1.98 - 5.93) | 97 | TRUE | 7.29E-01 | UNCERTAIN |
| Nucleus Accumbens | *RTEL1 Splice Junction 32* | Non-GB | Wald ratio | 1 | 2.32E-10 | 5.92 (3.41 - 10.25) | 99 | TRUE | 8.16E-01 | UNCERTAIN |
| Putamen | *RTEL1 Splice Junction 32* | Non-GB | Wald ratio | 1 | 1.56E-10 | 5.35 (3.2 - 8.93) | 100 | TRUE | 7.66E-01 | UNCERTAIN |
| Substantia Nigra | *SLC8A1 Splice Junction 7* | All | Wald ratio | 1 | 5.72E-06 | 0.37 (0.24 - 0.56) | 98 | TRUE | 7.01E-01 | UNCERTAIN |

Results presented here are those which were deemed the most robust, i.e., met the Bonferroni-correct P value threshold ( $1.11 \times 10^{-5}$ ) for the MR analysis, had
strong evidence of colocalisation ( $H4 > 80\%$ ) and correctly orientated Steiger filtering direction.

Three genes (*FAIM, PHLDB1, RTEL1*) had robust evidence of a causal effect on glioma risk within multiple tissues, with different lead SNPs, but all were in high LD (R^2^ > 0.7). ^30^ One gene (*CEP192*) with robust evidence of a causal effect on GB, whereas all other genes (*FAIM, HEATR3, PHLDB1, SLC8A1* and *RTEL1*) had a causal effect on all glioma, or more than one outcome tested (Table 2).

In summary, we found six genes for which there was robust evidence that splicing had a causal effect on at least one glioma subtype in at least one brain region (24 causal effects in total, Figure 4). All results for all sQTL sensitivity analyses are described in Supplementary Table S1.

**Figure 4.**
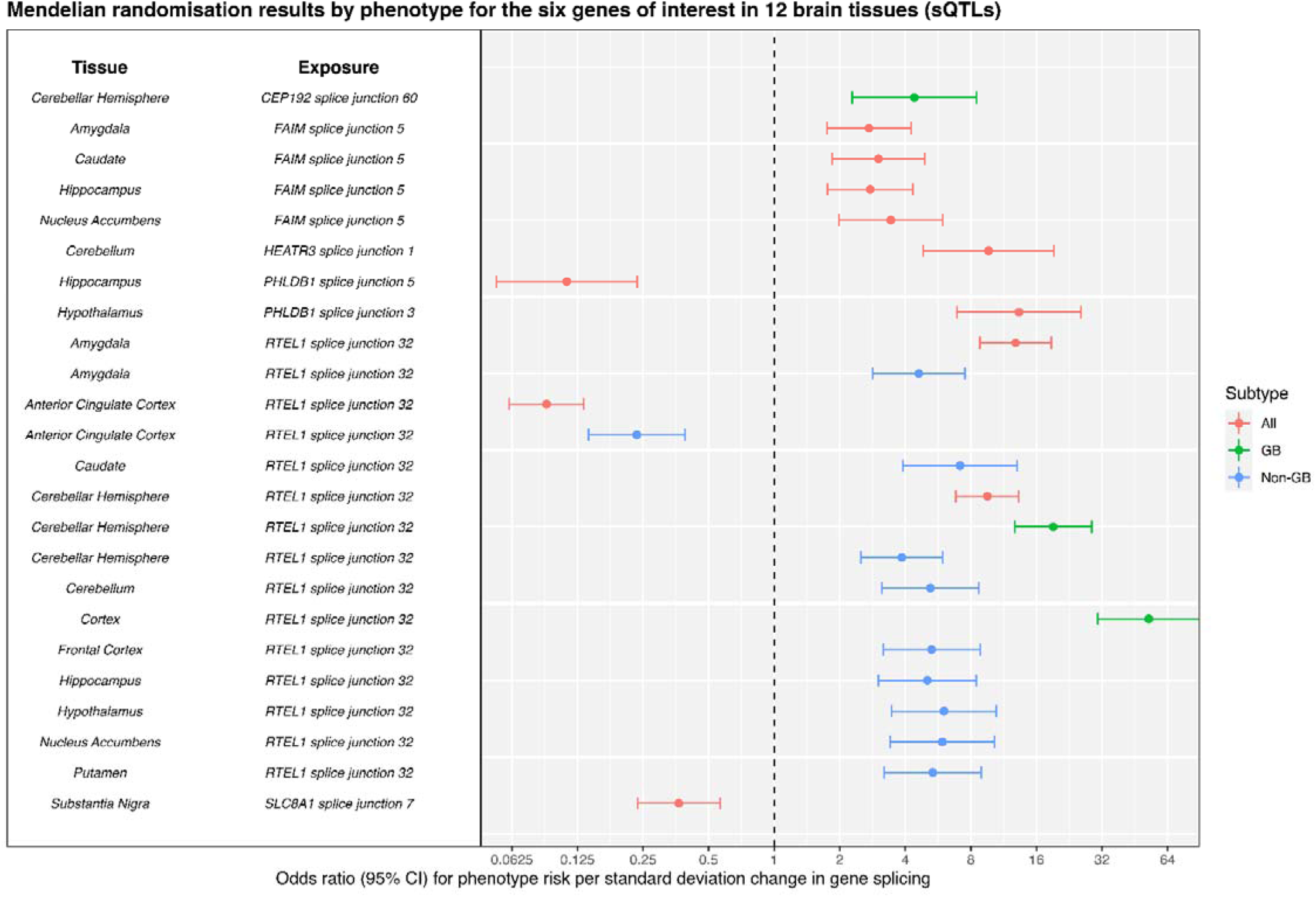
Forest plot showing Mendelian randomisation results by subtype (all glioma, GB and non-GB) for the six genes of interest in 12 brain tissues in the gene-splicing analysis. Results have strong evidence of colocalization (H_4_ >= 80%) and correct Steiger directionality.

### Estimating the causal effects of protein abundance on glioma risk

Of the 618 proteins that could be instrumented, all (100%) had the relevant SNPs available in the glioma GWAS are therefore MR could be performed. We only found one protein with evidence for a causal effect on GB in DLPFC (P < 8.09 x 10^-5^), SCFD1 (OR 0.17 (95%CI 0.07 to 0.40); *P* = 6.67 x 10^-5^), Supplementary Figure S3).

We applied the same sensitivity pipeline but there was not sufficient SNPs in the *cis*-region of SCFD1 to perform colocalisation. We used LDCheck (see methods) to determine weak proxy evidence of colocalisation. However, none of the SCFD1 SNPs in the glioma dataset reached our most lenient threshold of P < 5 x 10^-6^. Therefore, we could not conclude that this pQTL had sufficient causal evidence to be associated with glioma.

In summary, there were no proteins with robust causal evidence from the MR and sensitivity analyses. All results for causal pQTL sensitivity analysis are described in Supplementary Table S1.

### Novel genes

From our multi-omic analysis, we found robust evidence for four genes that not been implicated with glioma risk in previous genetic studies: *CEP192* (18p11.21), *FAIM* (3q22.3), *HBEGF* (5q13.3) and *SLC8A1* (2p22.1).

We found that increased *HBEGF* expression in the cortex is causal for all glioma (OR 1.36 (95%CI 1.19 to 1.55); P = 4.41 x 10^-6^). We found there was also increased risk of glioma in the GB and non-GB subtypes in the cortex, but they did not meet our Bonferroni-corrected P value threshold (P = 5.53 x 10^-6^ and 2.01 x 10^-2^, respectively) (Table 1, Figure 5).

**Figure 5.**
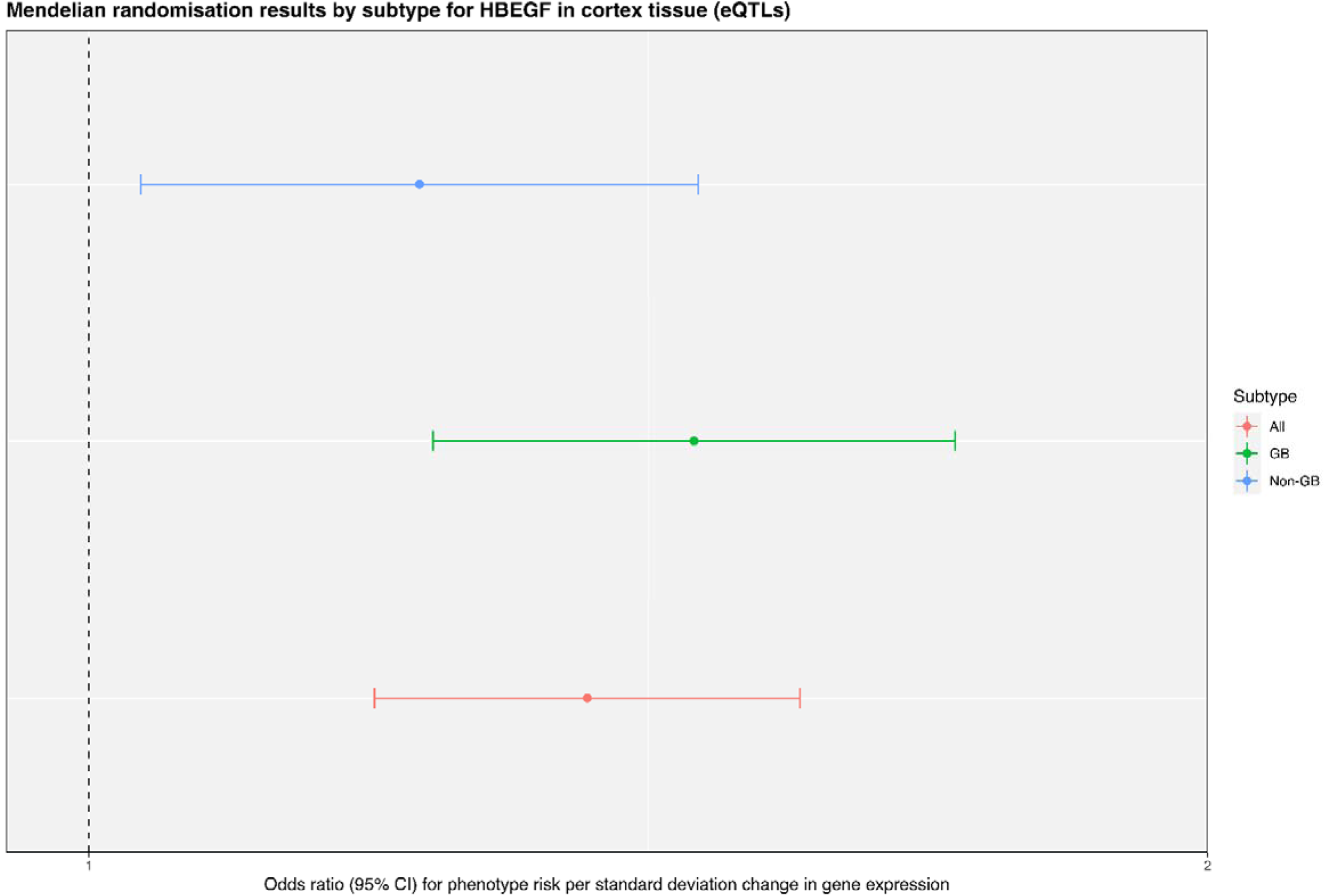
Forest plot showing Mendelian randomisation results by subtype (all glioma, GB and non-GB) for HBEGF in cortex tissue in the gene expression analysis. All subtypes show an increased risk of developing glioma, but only all glioma surpassed our Bonferroni-corrected P value threshold (P = 5.00 x 10^-6^).

We found that *FAIM* isoforms in four tissues were causal for all glioma (OR 2.72 to 3.43; P = 1.03 x 10^-5^ to 1.09 x 10^-5^). There was also an increased risk of glioma in the GB and non-GB subtypes in these tissues, but they failed to meet our Bonferroni-corrected P value threshold (P = 4.36 x 10^-5^ to 9.64 x 10^-3^). We found that *CEP192* splicing in the cerebellar hemisphere is causal for GB (OR 4.40 (95%CI 2.28 to 8.48; P = 9.78 x 10^-6^). There was also an increased risk of glioma in all glioma (but not non-GB), but this failed to meet the Bonferroni-corrected P value (P = 1.74 x 10^-3^) (Table 2, Figure 6). We found that *SLC8A1* splicing in the substantia nigra was protective for all glioma (OR 0.37 (95%CI 0.24 to 0.56); P = 5.72 x 10^-6^). There was also a decreased risk of glioma in the GB and non-GB subtypes in this tissue, but they failed to meet our Bonferroni-corrected P value threshold (P = 5.67 x 10^-4^ and 1.81 x 10^-4^, respectively) (Table 2, Figure 6).

**Figure 6.**
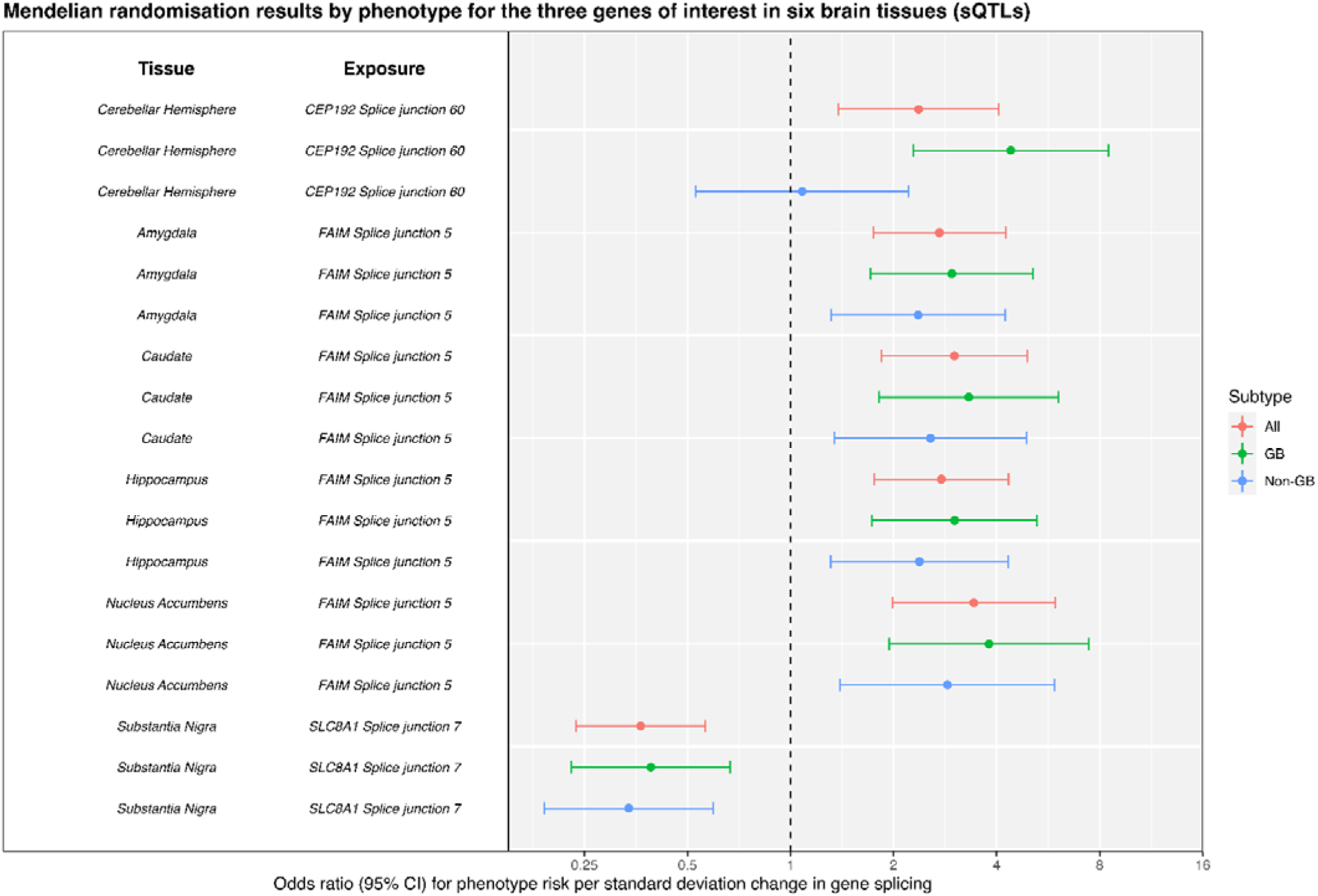
Forest plot showing Mendelian randomisation results by subtype (all, GB and non-GB) for CEP192, FAIM and SLC8A1 in six tissues in the gene-splicing analysis. All subtypes showed the same direction of effect on glioma risk within tissues, but not all passed our Bonferroni-corrected P value threshold (P = 1.11 x 10^-5^)

### Overlap in signals between molecular traits

We analysed all 16 robust molecular traits that we identified in our analysis in each tissue type across all three datasets, regardless of P value. We performed 104 tests, of which 82 molecular traits had passed the Bonferroni corrected P value in the primary analysis. Of the 22 other molecular traits, 12 passed the more lenient Bonferroni-corrected P value (P = 4.81 x 10^-4^), of which only *FAIM* in the cerebellum in all glioma (P = 6.93 x 10^-5^) showed potential overlap between molecular traits (gene expression and alternative splicing). Additionally, we found that the effect of *HEATR3* appears to be eQTL-specific (P = 1.95 x 10^-10^ to 6.74 x 10^-12^) despite finding a robust causal effect in the alternative splicing analysis (P = 1.29 x 10^-10^, Supplementary Table S2). The causal effect is likely driven through expression of the canonical gene isoform, which is incorrectly measured as an alternative splice isoform due to the way in which eQTL and sQTL are measured (see discussion).

### Specificity analysis

To annotate the potential causal pathways between the robust molecular traits and glioma, we used PheWAS data, and known glioma risk factors with pre-existing evidence. *HBEGF* is commonly linked with other cancer types so we wanted to explore this putative oncogene further. ^44^ Similarly, *RTEL1* is linked with telomere length which has been previously implicated in observational and MR studies to have an association with glioma risk. Additionally, the considerable number of tissues in which we found a causal effect warranted further analyses. ^45–47^

We used the IEU OpenGWAS project PheWAS tool and PhenoScanner v2 to conduct a broad search for phenotypes which were strongly associated with the lead SNP(s) for *HBEGF* (rs4150197) and *RTEL1* (rs6062302, rs2297440, rs2315009, rs3208007, rs2777941) at genome-wide significance (P < 5 x 10^-8^). ^31,33^ We found that rs4150197 was associated with anthropometric traits such as hip circumference and weight, and various image-derived phenotypes from diffuse MRI (Supplementary Table S2). We found that rs6062302 and rs2297440 were already associated with glioblastoma (P = 1.00 x 10^-13^ and P = 4.00 x 10^-46^, respectively) and rs2297440 was associated with glioma (P = 2.00 x 10^-42^). We also found significant associations with phenotypes associated with the immune system such as dermatitis, eczema, Crohn’s disease, inflammatory bowel disease and allergic rhinitis (Supplementary Table S2).

For the 16 molecular traits that had strong evidence for a causal effect on glioma, we performed a follow-up ad hoc analysis to determine whether these molecular traits also had causal evidence with putative glioma risk factors. We found that of the molecular traits with robust causal evidence, only *HBEGF* showed no evidence for non-specific effects with any of the traits we identified. The trait that was associated with the most potential glioma genes was LDLc, which was associated (P < 0.05) with seven genes (*CDKN2B, FAIM, GALNT6, HEATR3, PHLDB1, PICK1, RTEL1*). (Supplementary Table S2).

### Differential expression in molecular traits

We searched for differential expression of *HBEGF* using the Rembrandt and TCGA studies, found in the GlioVis tool. We found no significant differences (P < 0.05) between non-tumour samples and any of the glioma subtypes but did find differential expression between GB and non-GB subtypes (P = 3.10 x 10^-5^ to 1.90 x 10^-10^) (Supplementary Figure S4A&B). ^48^

Additionally, we searched for differences in isoform expression of *RTEL1* using the GTEx portal tool. ^22^ We found that isoform expression was significantly lower between wild-type (CC) and the homozygous variant (TT) in our lead SNPs (P = 6.64 x 10^-18^ to 4.23 x 10^-75^). We found that the cerebellum had the highest isoform (ENST00000425905.5) expression (3.17 transcripts per million (TPM)), one of the tissues in which we found our results, followed by the cerebellar hemisphere (2.96 TPM) and testes (1.31 TPM). ^22^

### Annotation of ‘druggability’

We used OpenTargets and DGIdb to gather evidence of ‘druggability’ for molecular traits with robust evidence. ^36–38^ We found that 4/10 (40%) of the molecular traits identified in the expression analysis, and none of the molecular traits identified in the gene-splicing analysis were categorised as being part of the ‘druggable genome’. One novel result, *HBEGF,* had three drug-gene interactions (cetuximab, panitumumab and KHK-2866), the latter being an anti-HBEGF antibody which was trialled for use in advanced solid tumours and ovarian cancer. However, the study was discontinued after 22 months due to patients experiencing reversible neuropsychiatric toxicity, although the aetiology was not understood (Supplementary Table S2). ^49^

## Discussion

In this study, we used a combined MR-colocalisation framework to estimate the causal effect of genetically proxied gene expression, alternative splicing and protein abundance levels on genetic liability of glioma risk. We found robust evidence that causally implicated 16 molecular traits with glioma risk: ten gene expression evidence levels and six alternative splicing events (one gene shared both expression and splicing causal effects on glioma). Assessing different ‘omics’ data sources can be beneficial to understand the development of complex traits and for drug target-related analyses; each QTL provides an insight into different molecular processes. ^16,17^

For 11 of the genes that we found robust evidence for here, the loci have been previously implicated in GWAS/TWAS of glioma risk. ^9,10^ However, these studies did not establish causality of all these genes, as we have. Additionally, we found four genes that not been implicated in previous genetic studies: *CEP192* (18p11.21), *FAIM* (3q22.3), *HBEGF* (5q13.3) and *SLC8A1* (2p22.1). For brevity, we discuss *RTEL1* and a novel result, *HBEGF*.

*RTEL1* (Regulator of telomere elongation helicase 1) is a gene which encodes for a DNA helicase responsible for the elongation of telomeres, and is known to interact with the telosome complex, a group of proteins which protect the telomere caps of DNA. ^50^ *RTEL1* has previously been identified as a risk factor for glioma, and studies have tried to identify whether telomere length has an established causative effect on the risk of glioma, although no definitive conclusion has been reached.^12,14, 45–47^ *RTEL1*-associated sQTLs were associated with an exon-skipping event (GTEx intron ID: 63689132:63689750:clu_27064) present only in *RTEL1* transcript ENST00000425905 (P = 4.23 x 10^-75^ to 6.64 x 10^-18^). Therefore, it may be the case that previous conflicting evidence on the directionality of the effect of *RTEL1* expression on glioma risk may be confounded due to non-canonical transcript variants; though, further studies would be required to ascertain if this is the case. Furthermore, we observed that *RTEL1* sQTLs rs6063202 and rs2297440, were found to be strongly associated to glioblastoma (P = 1.00 x 10^-13^ and 4.00 x 10^-46^, respectively) and with allergy-related traits (P = 4.78 x 10^-8^ to 7.84 x 10^-19^, Supplementary Table S2). These results beget further study and could help to understand the links between allergies, glioma risk and *RTEL1*.

*HBEGF* (Heparin-binding epidermal growth factor) is a member of the EGF family of growth factors which is involved in the MAPK, STAT and PI3K/AKT pathways. ^44,51^ These molecular pathways have roles in regulating biological processes, such as positive regulation of cell growth, migration, and proliferation. ^52^ The role of *HBEGF* in the CNS is well characterised: it is highly expressed throughout; has roles in both the developing and adult brain, with studies indicating *HBEGF* contributes to glial and stem cell proliferation and is a ligand for EGFR.^53,54^ Furthermore, expression of *HBEGF* has been shown to be increased significantly in many human cancer types, and *HBEGF* mRNA expression has been shown to be increased two- to five-fold in glioblastoma cell lines compared to control brain tissue. ^55^ We found no significant differences between control and any glioma subtypes but did find differential expression between GB and non-GB subtypes in two independent datasets (Supplementary Figure S4A&B). ^48,56^ We also found that the instrumented *HBEGF* eQTL (rs4150197) had no associations with cancer risk or any putative risk factors for glioma (Supplementary Table S2). Unlike *RTEL1*, we observed no evidence for an effect through non-canonical transcripts or proteins. Altogether, our causal evidence implicates the expression of *HBEGF* in the brain as a potentially novel biomarker, though more research would be required to ascertain its role. Given that there are drugs which target *HBEGF*, glioma and *HBEGF* may make for an interesting target-indication pair for future studies.

### Strengths

We aimed to identify molecular traits with the most robust evidence for causal roles in glioma risk, by combining strong MR evidence with colocalisation and Steiger filtering. To test if the causal effects were driven via putative glioma risk factors, we tested for potential pleiotropic effects in known risk factors for glioma: allergies, asthma, altered low-density lipoprotein cholesterol (LDLc) levels and alcohol consumption. This may provide evidence of horizontal pleiotropy, which may make it harder to draw decisive conclusions of causality ^27,29^ However, this would require follow up studies to investigate which could lead to potentially elucidating how these putative risk factors and molecular traits are linked to glioma.

### Limitations

Despite using relatively large datasets, our analyses are still likely to suffer from limited statistical power due to restricted sample sizes, particularly in the sQTL dataset (n = 114 to 209). This might lead to some important causal effects not being identified. All three QTL datasets were derived from a mixture of case and control samples. However, the cases were derived from individuals whose disease (e.g. Alzheimer’s disease in BrainQTL) have no known link to glioma risk.

Additionally, as we used the GTEx sGenes, only the top splice event per gene was measured, and therefore there were many splice events which were not tested in our MR analysis.

Most of the MR analyses used a single SNP instrument, which restricts the type of sensitivity analyses that could be performed; however, this is a common phenomenon observed when conducting MR with molecular traits. ^29^

SNPs can act via multiple molecular QTL pathways and are not mutually exclusive; SNPs affecting gene expression can also be associated with alternative gene splicing of the same genes. As eQTL and sQTL are both measured by quantifying mRNA levels, this was not unexpected. ^57^ These SNPs may be legitimately affecting both gene expression and alternative splicing, however if a particular splice variant alters the ability of a gene to be efficiently measured, it appears that gene expression is altered, and the effect is driven by the splicing event. ^58^ Furthermore, some probes for mRNA will detect commonly splice variants as canonical transcripts, which are included in ‘bulk tissue’ eQTL analysis. This can lead to the false assumption that both expression and splicing events are driving the causal effect, when this is not the case. ^59^

Our analysis is limited to individuals with European ancestry; therefore, it will be important to extend these analyses to individuals of alternative ancestries as such data becomes available. ^60^ Previous GWAS and observational studies in non-European cohorts have found novel associations which we did not find in our study e.g., *GSTP1*, which may illustrate that the loci appear to be population specific, or may be observational, and not causal. ^61,62^

The nature of glioma can result in variation in inter-patient, intra-tumoural, subtype and spatiotemporal heterogeneity, which can lead to variation in tissue recovery at biopsy. ^63,64^ These factors can result in deviation in the profile of disease, which can make our results more generalisable and less specific to one subtype of glioma. To make our analysis more precise, we would require more granular tissue and cell-specific data.

## Conclusion

We conducted a robust multi-omic causal analysis for gene and protein molecular traits in brain tissue on glioma risk. We used tissue-specific molecular data and glioma subtype information to explore the nature of the causal relationships identified. Here, we combined MR with colocalisation and Steiger filtering to ensure the robustness of our results.

We provide robust multi-omic causal evidence for 11 previously implicated genes which affect the risk of glioma. We also present novel evidence for a causal effect of increased *HBEGF* expression in cortex on increased risk of all glioma, and present novel evidence for causal effects of *CEP192*, *FAIM* and *SLC8A1* splicing in multiple tissues on variable risk of all glioma and GB. Additionally, we show evidence for the causal effect of increased *HEATR3* expression and alternative splicing on increased risk of all glioma, although we propose that this effect is eQTL-specific.

We did not identify any causal proteins for glioma risk, likely due to the limited sample size of the currently available brain pQTL data.

We focussed on the causal effects of increased *HBEGF* expression and alternative splicing of *RTEL1* and use multiple investigate methods to provide evidence to understand the biological mechanisms that these genes may play in the risk of glioma.

## Abbreviation List

CNS: Central nervous system
DGIdb: Drug gene interaction database
eQTL: Expression quantitative trait loci
GB: Glioblastoma
GWAS: Genome-wide association study
LD: Linkage disequilibrium
LDLc: Low-density lipoprotein cholesterol
MR: Mendelian randomisation
OR: Odds ratio
pQTL: Protein quantitative trait loci
QTL: Quantitative trait loci
sQTL: Splicing quantitative trait loci
TPM: Transcripts per Million
WHO: World Health Organisation

## Declarations

### Consent for publication

Not applicable

### Availability of data and materials

The MetaBrain meta-analyses eQTL data can be accessed at https://www.metabrain.nl/. The GTEx sQTL v8 data can be accessed at https://gtexportal.org/home/datasets. The BrainQTL pQTL data can be found at https://www.synapse.org/#!Synapse:syn24172458.The glioma data may be accessed under the European Genome-phenome Archive accession number EGAD00010001657 (https://www.ebi.ac.uk/ega/datasets/EGAD00010001657).

### Competing interests

The authors declare they have no competing interests.

### Funding

ZAT, LJA, LP and JWR receive support from the UK Medical Research Council Integrative Epidemiology Unit at the University of Bristol (MC_UU_00011/4, MC_UU_00011/1). ZAT receives funding from Southmead Hospital Charitable Funds: Brain tumour bank and research fund 8036. LJA and KMK are funded by Cancer Research UK (grant number C30758/A29791). JWR received funding from Biogen for unrelated projects. KMK is funded by Innovate (grant number 10027624). This study was supported by the National Institute for Health and Care Research Bristol Biomedical Research Centre (NIHR203315) and Cancer Research UK (grant number C18281/A29019 and C18281/A30905). The views expressed are those of the author(s) and not necessarily those of the NIHR or the Department of Health and Social Care.

### Authors’ contributions

ZAT wrote the manuscript, performed all statistical analysis and interpreted results; LJA and HZ revised the manuscript; JZ advised on statistical methodology; LP substantially revised the manuscript and assisted with statistical analysis; JWR substantially revised the manuscript, assisted with statistical analysis and interpreted results and designed the study; KMK designed the study. All authors have approved the submitted version of the manuscript.

## Data Availability

All data produced in the present study are available upon reasonable request to the authors.

https://www.metabrain.nl/

https://gtexportal.org/home/datasets

https://www.synapse.org/#!Synapse:syn24172458

## Acknowledgements

We would like to acknowledge all patients who provided data that made this work possible.

## Notes

### Competing Interest Statement

The authors have declared no competing interest.

### Author Declarations

The MetaBrain meta-analyses eQTL data can be accessed at https://www.metabrain.nl/. The GTEx sQTL v8 data can be accessed at https://gtexportal.org/home/datasets. The BrainQTL pQTL data can be found at https://www.synapse.org/#!Synapse:syn24172458. The NIAGADS pQTL data can be found at https://www.niagads.org/datasets/ng00102. The glioma data may be accessed under the European Genome-phenome Archive accession number EGAD00010001657 (https://www.ebi.ac.uk/ega/datasets/EGAD00010001657).

### Summary of Updates

Newer version of the MetaBrain data in analysis; New analysis workflow; Major revisions to the manuscript; Author list amended

