## Supplementary Figures for "Multi-omics Mendelian randomisation using expression, splicing and protein quantitative trait loci: identification of novel drug targets for gliomagenesis"

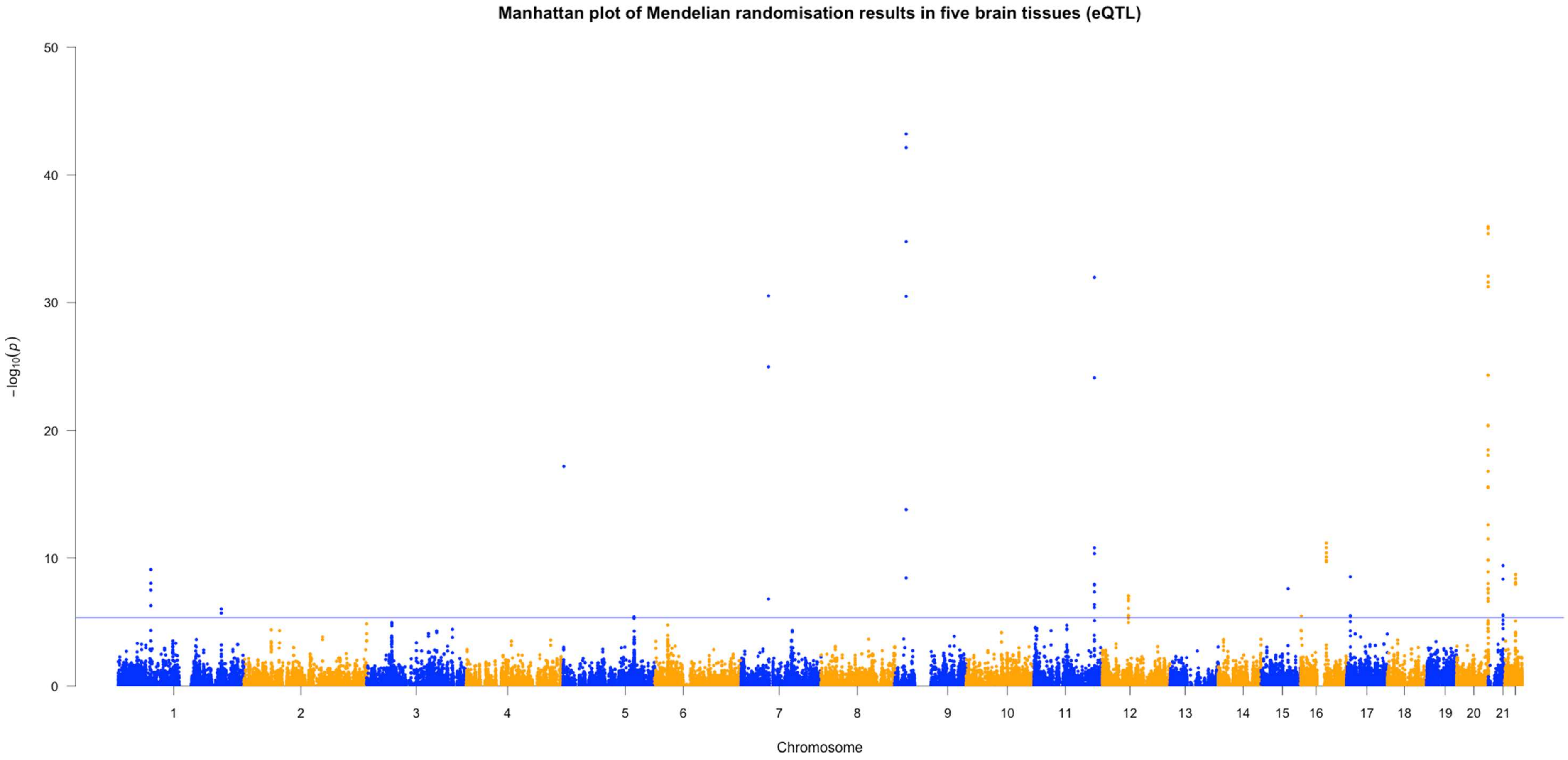


*Figure S1. Manhattan plot of Mendelian randomisation in five brain tissues in the gene expression analysis. The Bonferroni corrected P value was set at $P < 5.00 \times 10^{-6}$.*

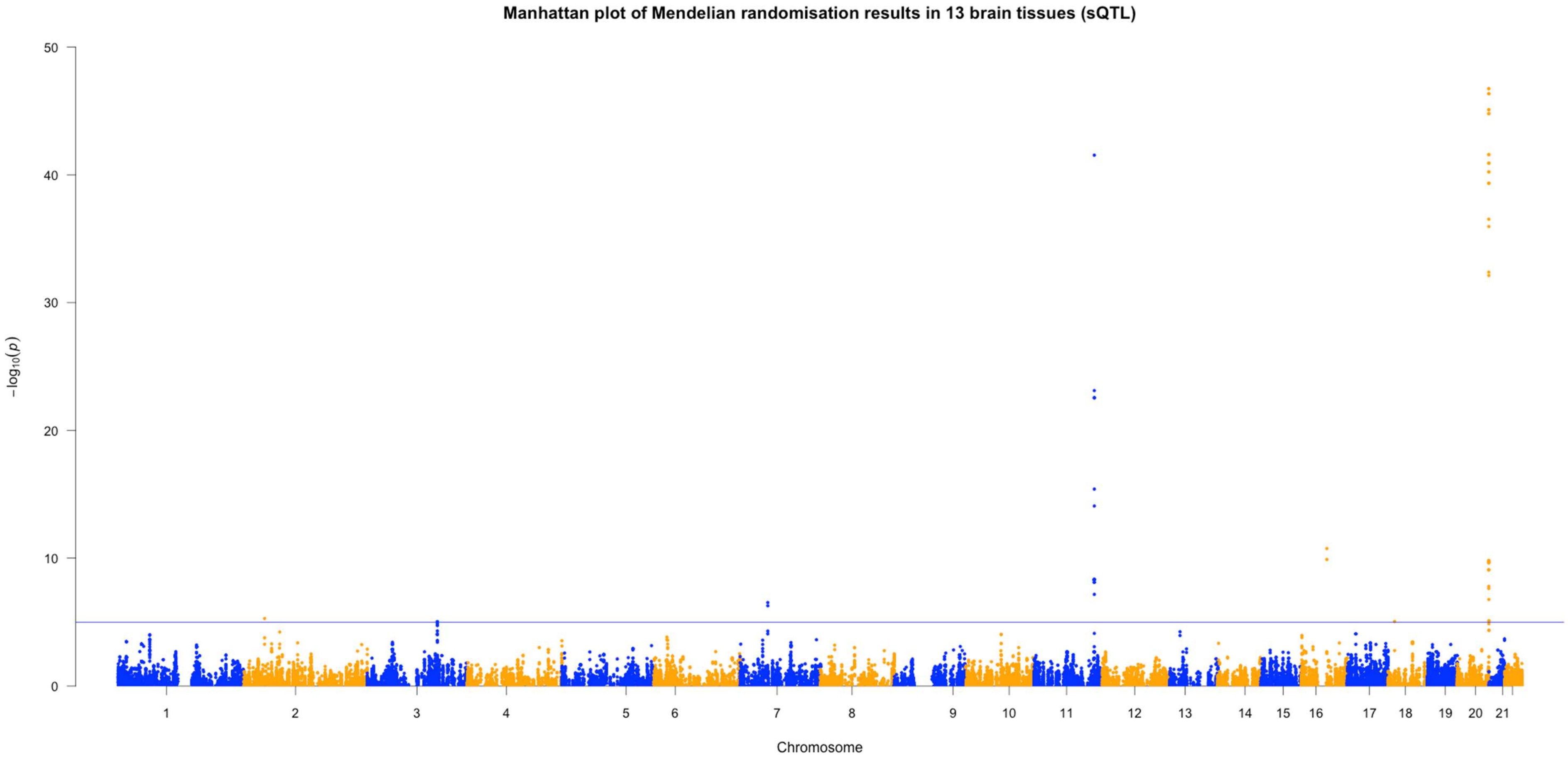


*Figure S2. Manhattan plot of Mendelian randomisation in 13 brain tissues in the gene splicing variation analysis. The Bonferroni corrected P value was set at $P < 1.11 \times 10^{-5}$.*

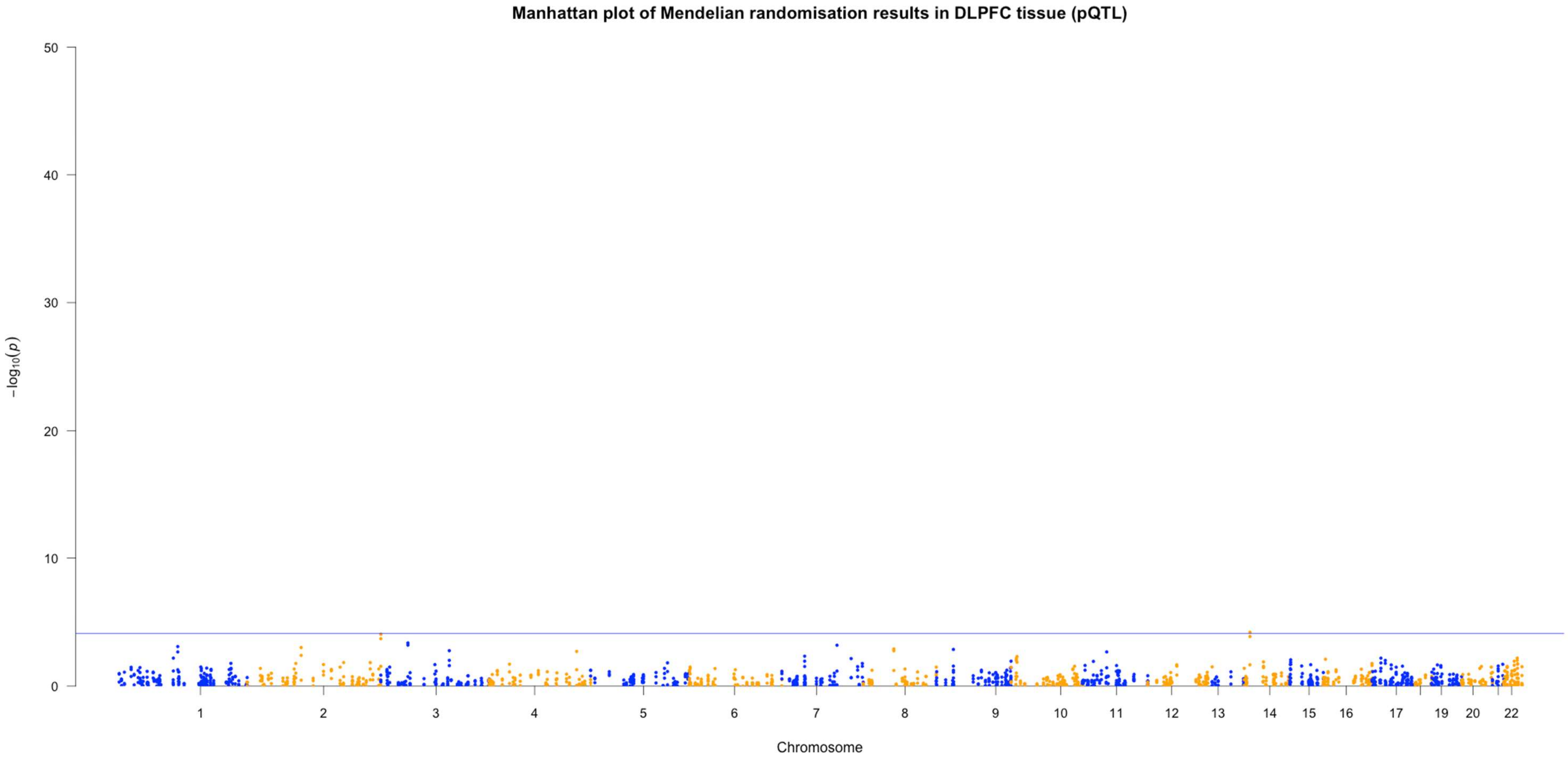


*Figure S3. Manhattan plot of Mendelian randomisation in dorsolateral pre-frontal cortex (DLPFC) in the protein abundance analysis. The Bonferroni corrected P value was set at $P < 8.09 \times 10^{-5}$.*

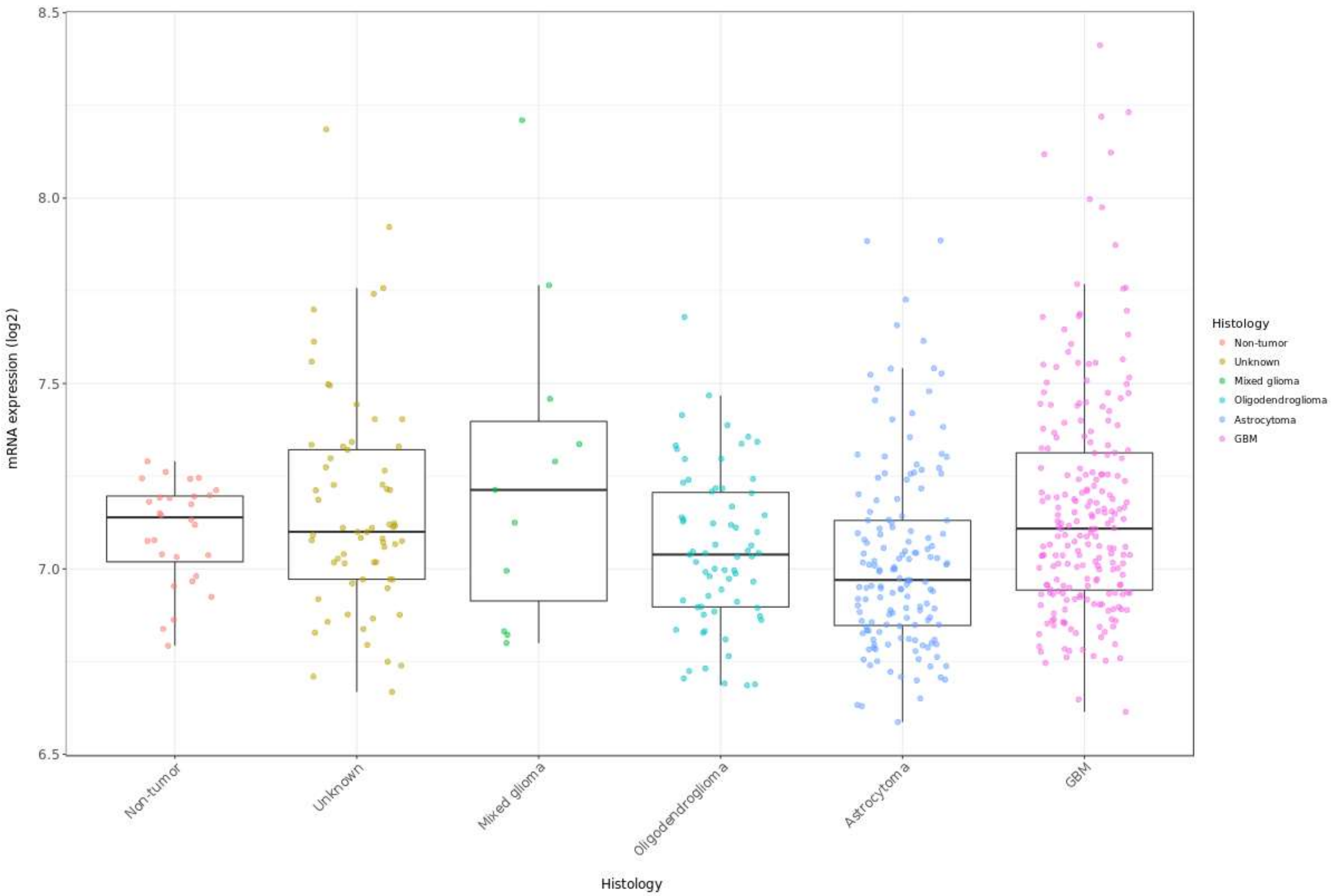


*Figure S4a. Box plot of HBEGF mRNA expression (log2) in 537 glioma cases (non-tumour (n=28), unknown (n=65), mixed glioma (n=11), oligodendroglioma (n=67), astrocytoma (n=147), GB (n=219) in the Rembrandt dataset. There was a significant difference ($P > 0.05$) in mRNA expression between GB and astrocytoma ($P = 3.10 \times 10^{-5}$) and unknown subtype and astrocytoma ($P = 0.01$).*

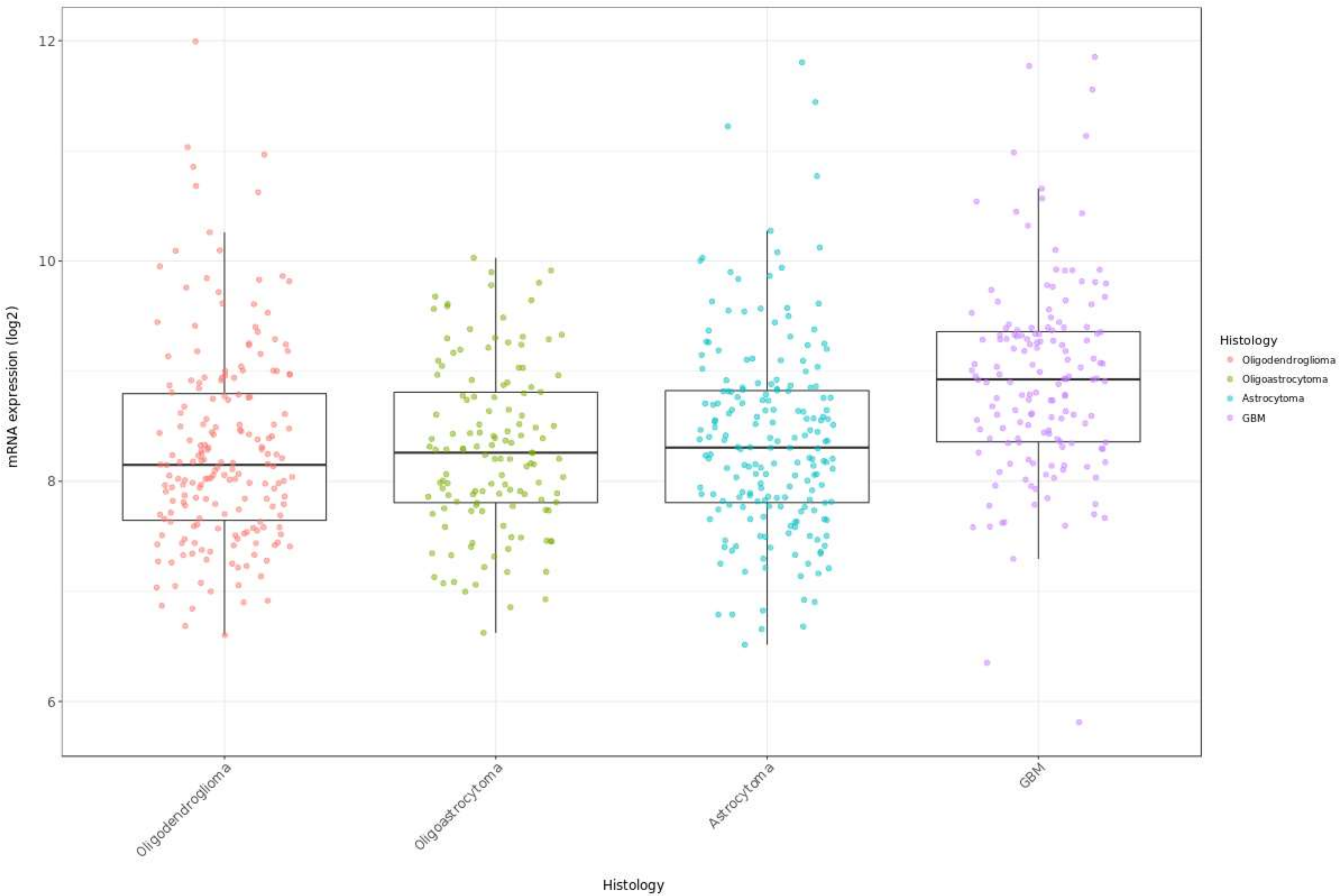


*Figure S4b. Box plot of HBEGF mRNA expression (log2) in 667 glioma cases (oligodendroglioma (n=191), oligoastrocytoma (n=130), astrocytoma (n=194) and GB (n=152)) in the TCGA cohort. There was a significant difference in mRNA expression between GB and oligodendroglioma (P = 1.90 x $10^{-10}$), GB and oligoastrocytoma (P = 1.00 x $10^{-8}$) and GB and astrocytoma (P = 3.50 x $10^{-8}$).*
